# Enhanced detection of neonatal invasive infection clusters in South Africa using epidemiological and genomic surveillance data

**DOI:** 10.1101/2025.11.10.25339895

**Authors:** Liliwe Shuping, Husna Ismail, Rudzani Mashau, Stanford Kwenda, Kathryn E. Holt, Rindidzani E. Magobo, Olga Perovic, Susan T. Meiring, Vanessa C. Quan, Nelesh P. Govender, Baby GERMS-SA

## Abstract

**Introduction:** Invasive bacterial infections, particularly those caused by the ESKAPE group, account for a substantial proportion of neonatal deaths in low- and middle-income countries, yet the contribution of healthcare-associated transmission of such infections remains poorly defined. Pathogen whole genome sequencing (WGS) offers superior resolution to delineate transmission events compared to conventional methods. We aimed to determine the proportion of neonatal infection cases attributable to healthcare-associated transmission by comparing classical epidemiological and enhanced genomic approaches.

**Methods:** We retrospectively analyzed infants with culture-confirmed bloodstream infection or meningitis from six lower-tier hospitals in South Africa from September 2019 through October 2020. Isolates including *Klebsiella pneumoniae*, *Acinetobacter baumannii*, *Staphylococcus aureus*, *Enterococcus faecium*, *Enterococcus faecalis*, and *Escherichia coli* underwent antimicrobial susceptibility testing (AST) and WGS. Clusters were delineated using: 1) SNP-EPI (WGS-based reference standard), defined as 2 or more cases in the same hospital within 14 days and 11 or less single nucleotide polymorphisms (SNPs) (10 or less for *A. baumannii*); and 2) AST-EPI (conventional approach), defined as 2 or more cases with identical AST patterns within 14 days in the same hospital. The healthcare-associated transmission rate was calculated as the proportion of all cases allocated to SNP-EPI clusters.

**Results:** We included 428 cases of infection with both AST and WGS data, including *K. pneumoniae* (40%, n=172), *A. baumannii* (23%, n=98), and *S. aureus* (21%, n=90). The overall healthcare-associated transmission rate was 31% (131/428 cases); this was highest for *A. baumannii* (50%, 47/98) followed by *K. pneumoniae* (42%, 72/172) and *S. aureus* (11%, 10/90), with substantial variation by facility (range, 5%-95%). We identified 39 SNP-EPI clusters across all hospitals, mostly involving multi-drug-resistant (MDR) strains, including large outbreaks of carbapenem-resistant *K. pneumoniae* ST152 and high-risk *A. baumannii* ST1 and ST2 clones. The AST-EPI criteria detected only 28 clusters and yielded a lower overall healthcare-associated transmission rate of 18%. Compared to the SNP-EPI approach, the AST-EPI method particularly underestimated transmission for *K. pneumoniae* (23% vs. 42%) and *A. baumannii* (29% vs. 48%), often fragmenting larger, persistent SNP-EPI clusters.

**Conclusions:** Healthcare-associated transmission contributed substantially to the burden of invasive bacterial infections among South African infants, primarily driven by MDR *K. pneumoniae* and *A. baumannii*. WGS provided greater resolution than conventional methods for cluster detection. Genomic surveillance should be integrated into routine practice for effective outbreak detection and informed infection prevention and control responses in low-resource settings.

## Introduction

Invasive infections account for 17%-29% of neonatal deaths in low- and middle-income countries (LMICs) [1]. A large proportion of this infection incidence is probably related to healthcare-associated transmission events, caused by inadequate infection prevention and control (IPC), during which pathogens are transferred to patients either from a contaminated environment or on the hands of healthcare workers. Unfortunately, many of these healthcare-associated infections are caused by the ESKAPE pathogens (i.e. *Enterococcus faecium*, *Staphylococcus aureus*, *Klebsiella pneumoniae*, *Acinetobacter baumannii*, *Pseudomonas aeruginosa*, *Enterobacter* species), which are commonly resistant to multiple antimicrobial agents. In South Africa, the incidence of culture-confirmed blood and central nervous system infections in the newborn population is 6.0 per 1000 live births (95% confidence interval [CI], 6.0-6.1), with ESKAPE pathogens *Klebsiella pneumoniae*, *Acinetobacter baumannii* and *Staphylococcus aureus*, accounting for 50% of these infections [2].

Furthermore, healthcare transmission events probably result in infection outbreaks which are undetected owing to insufficient surveillance infrastructure. An epidemiological approach, in which temporal and spatial linkage of events with or without pathogen antimicrobial susceptibility profiles are used to detect outbreaks, is not always adequate [3]. Healthcare infection surveillance incorporating pathogen whole genome sequencing (WGS) is currently the most robust approach, yielding granular pathogen data to characterize transmission pathways and detect healthcare-associated outbreaks [3, 4]. In a multi-center surveillance study, the combination of genomic data and epidemiological rule-based thresholds provided sufficient information to prospectively detect clusters of multi-drug-resistant (MDR) pathogens that would otherwise have been missed by pathogen antimicrobial susceptibility and epidemiological links alone [4, 5]. A recent study demonstrated that incorporating prospective WGS data resulted in early detection of clusters, rapid action, and aversion of large outbreaks [6]. Prevention of infections was associated with a net US$695,706 cost reduction over two years.

The implementation of genomic surveillance in LMICs has the potential to reduce the burden of infection by rapidly detecting outbreaks, identifying infection sources and informing targeted IPC interventions. However, the added value of genomic data to IPC activities in low-resource settings has not been demonstrated, in part due to a lack of setting-specific studies. We conducted a study across six neonatal units in South Africa, integrating WGS, epidemiological data, and antimicrobial susceptibility test (AST) results. Our primary objective was to estimate the proportion of infection cases linked to healthcare-associated transmission by comparing a traditional epidemiological approach with an enhanced genomic approach. We aimed to demonstrate the potential value of genomic data for the timely detection of outbreaks during continuous real-time surveillance, as well as for monitoring transmission dynamics and assessing the impact of IPC interventions through routine retrospective or periodic data reviews.

## Methods

### Baby GERMS-SA surveillance

In the Baby GERMS-SA surveillance, infants with culture-confirmed bloodstream infection or meningitis were retrospectively enrolled between September 2019 and October 2021 at six lower-tier hospitals in South Africa [7]. The surveillance included babies admitted to a neonatal unit or those aged ≤6 months admitted to a pediatric unit who had a pathogen isolated in culture from blood or cerebrospinal fluid. Demographic and clinical information of the case-patients was abstracted by medical officers from retrospectively-scanned medical records. National Health Laboratory Service (NHLS) microbiology laboratories at sentinel hospitals submitted isolates from case-patients to reference laboratories at the National Institute for Communicable Diseases (NICD) for species-level identification, AST and WGS. WGS was performed only on the first isolate of each case-patient with infection due to *Enterococcus faecalis*, *Enterococcus faecium*, *S. aureus*, *K. pneumoniae*, *A. baumannii*, and *Escherichia coli*. The Baby GERMS-SA study received approval from the Human Research Ethics Committee of the University of the Witwatersrand (M190320). Informed consent was waived because the study involved the retrospective collection of clinical data through medical record review after discharge from hospital or death. No personal identifiers were collected, and each patient was assigned a unique study ID.

### Reference laboratory testing

Bacterial strains were transported on Dorset transport medium (Diagnostic Media Products, NHLS, Johannesburg, South Africa). The species of viable isolates was confirmed using matrix-assisted laser desorption/ionization time-of-flight mass spectrometry (MALDI-TOF MS) (Bruker Daltonik, Bremen, Germany). Minimum inhibitory concentrations (MICs) were determined using the MicroScan WalkAway 7465 instrument (Beckman Coulter, Sacramento, USA) and MIC values were interpreted according to Clinical and Laboratory Standards Institute (CLSI) recommendations (M100, 31st edition, 2021).

### Whole genome sequencing

DNA was extracted from overnight broth cultures using the QIAamp DNA Micro Kit (Qiagen, USA), following the manufacturer’s protocol. The DNA concentration was measured using a Nanodrop 2000 spectrophotometer (Thermo Fisher Scientific, UK), and the DNA purity was assessed using a Qubit 4 fluorometer (Thermo Fisher Scientific). The Illumina DNA Prep kit (Illumina, San Diego, USA) was used to generate multiplexed, paired-end libraries of 2x 150 bp. Sequencing was performed on the Illumina NextSeq 550 platform, targeting 100x coverage per isolate.

### Bioinformatic analysis

Raw paired-end sequencing reads were processed using the Jekesa pipeline v1.0 for read quality control, species identification, and multi-locus sequence typing [8]. Resistance genes were identified by scanning assembled contigs with AMRFinderPlus [9], ABRicate, and staramr against PointFinder, ResFinder [10], and AMRFinderPlus databases, with outputs summarised using hAMRonization [11]. For phylogenetic analysis, assembled genomes were aligned to reference genomes (*K. pneumoniae* HS11286, *A. baumannii* ATCC17978, and *S. aureus* Newman) using scapper [12], with recombinant regions removed using Gubbins v3.2.1. Variable sites were extracted using snp-sites v2.5.1, and maximum-likelihood phylogenetic trees were generated using IQ-TREE v2.0.3 with GTR + F + ASC + R4 model and 1000 UFBoot2 bootstrap approximations. Trees were visualised using iTOL (v7). Pairwise single nucleotide polymorphism (SNP) distances calculated with snp-dists v0.8.2 were used for cluster analysis with SNP2Cluster v0.5.4 [13].

### Cluster analysis

We used the Baby GERMS-SA epidemiological data, reference laboratory AST data, and WGS data to retrospectively identify clusters and/or outbreaks of *E. faecalis*, *E. faecium*, *S. aureus*, *K. pneumoniae*, *A. baumannii*, and *E. coli*. We defined cases according to the main study [7]. Bloodstream infection or meningitis was defined as a pathogenic organism isolated from a blood or cerebrospinal fluid culture, respectively. For the purpose of this analysis, we considered patients with more than one pathogen as having separate episodes. If the same pathogen was isolated >14 days after an initial culture-positive specimen, the patient was considered to have a new infection episode. Cases of *P. aeruginosa* and *E. cloacae* complex were not included due to a lack of WGS data (**Fig S1**).

### SNP-EPI and AST-EPI clusters

Clusters were detected using two approaches: the reference standard approach, in which we used WGS data to ascertain the relatedness of isolates (i.e. SNP-EPI clusters), and a conventional approach, in which pathogen AST patterns were used (i.e. AST-EPI clusters). A SNP-EPI cluster was defined as ≥2 cases in the same hospital with culture-positive specimens collected within 14 days of each other and a SNP cut-off of ≤11 (with the exception of *A. baumannii* for which a ≤10 SNP cut-off was used) [14–16]. AST-EPI clusters were defined as ≥2 cases with identical pathogen identification and AST patterns, with culture-positive specimens collected within 14 days of each other, and admission to the same hospital. We calculated the proportion of cases associated with healthcare transmission by dividing the number of cases allocated to clusters by all cases included in the cluster analysis, i.e. all those with both AST and WGS data available. Due to a wide range of adopted SNP cut-offs for isolate relatedness in outbreaks [14] we conducted sensitivity analyses for the primary SNP-EPI definition, varying SNP cut-offs and the time between culture-positive specimen collection dates, including combinations of ≤11 SNP-60 days, ≤20 SNP-14 days, ≤20 SNP-60 days and ≤25 SNP-45 days.

### Statistical analysis

Characteristics of case-patients were compared using Chi square test or Fisher’s exact test for categorical variables and the Kruskal Wallis test for continuous variables. We assessed the risk of death by causative pathogen using logistic regression. We explored factors associated with being part of a SNP-EPI cluster using a multivariable logistic regression. We included known infection risk factors such as age, prematurity, body weight at invasive specimen collection, and healthcare related factors such as prior antibiotic use, breastfeeding, and presence of medical devices, among others. Analysis was performed using Stata (v18.5, StataCorp LLC, Tx, USA) and R statistical software (v 5.4.1).

## Results

Of the 1079 cases of bloodstream infection (97%, n=1051) or meningitis (3%, n=28) (**Fig S1**), we included 428 (40% of all cases) with both AST and WGS data. *K. pneumoniae* (40%, n=172), *A. baumannii* (23%, n=98), and *S. aureus* infections (21%, n=90) accounted for most cases. The median age of these infants at diagnosis was 7 days (interquartile range [IQR]: 3-18) and 54% (n=221) were male (**Table 1**). The median age differed by causative pathogen, with *S. aureus* infected infants being older (9 days, IQR: 3-25, p=0.011). Of the 428 included infants, 22% (n=95) were in non-neonatal units at the time of specimen collection and a large proportion had late-onset sepsis (82%, 347/424). A majority of the infants weighed ≤1500 g (44%, 100/225), but most with *S. aureus* infections (72%, 28/39) weighed ≥2500 g while a majority with *K. pneumoniae* (45%, 44/97) and *A. baumannii* (68%, 38/56) weighed ≤1500 g (p<0.001). The overall mortality rate was 32% (88/277); this was higher in infants infected with *A. baumannii* (49%, 35/71) and *K. pneumoniae* (36%, 41/115) and lowest in infants with *S. aureus* infections (8%, 4/50) (p<0.001). Compared to *S. aureus* infections, the odds of death were 6 times higher in patients with *K. pneumoniae* infections (crude odds ratio [OR] 6.4, 95% CI 2.14-18.96, p=0.001) and 11 times higher in those with *A. baumannii* infections (OR 11.1, 95% CI 3.63-34.35, p<0.001) (**Table S1**).

**Table 1:** Characteristics of infant cases of bloodstream infection and meningitis, overall and by causative pathogen, at six sentinel hospitals in South Africa, September 2019-October 2020, n=428.

|  | Overall | *K. pneumoniae*<br>(172) | *A. baumannii*<br>(n=98) | *S. aureus*<br>(n=90) | *E. coli*<br>(n=27) | *E. faecalis*<br>(n=21) | *E. faecium*<br>(n=20) |  |
| --- | --- | --- | --- | --- | --- | --- | --- | --- |
|  | n (%) or median [IQR<sup>a</sup>] |  |  |  |  |  |  | p value |
| **Age (days)** | 7 [3 - 18] | 8 [5 - 18] | 6 [3 - 13] | 9 [3 - 25] | 4 [2 - 37] | 4 [1 - 10] | 4 [0 - 14] | 0.011 |
| **Sex** |  |  |  |  |  |  |  |  |
| Male | 221 (53.6) | 89 (54.9) | 50 (52.1) | 46 (52.9) | 19 (70.4) | 9 (45.0) | 8 (40.0) | 0.376 |
| **Hospital** |  |  |  |  |  |  |  |  |
| Hospital A | 97 (22.7) | 32 (18.6) | 23 (23.5) | 20 (22.2) | 9 (33.3) | 7 (33.3) | 6 (30.0) | <0.001 |
| Hospital B | 66 (15.4) | 31 (18.0) | 20 (20.4) | 7 (7.8) | 5 (18.5) | 2 (9.5) | 1 (5.0) |  |
| Hospital C | 65 (15.2) | 36 (20.9) | 7 (7.1) | 16 (17.8) | 2 (7.4) | 1 (4.8) | 3 (15.0) |  |
| Hospital D | 50 (11.7) | 29 (16.9) | 9 (9.2) | 6 (6.7) | 2 (7.4) | 1 (4.8) | 3 (15.0) |  |
| Hospital E | 95 (22.2) | 28 (16.3) | 29 (29.6) | 19 (21.1) | 8 (29.6) | 4 (19.0) | 7 (35.0) |  |
| Hospital F | 55 (12.9) | 16 (9.3) | 10 (10.2) | 22 (24.4) | 1 (3.7) | 6 (28.6) | 0 (0) |  |
| **Ward type** |  |  |  |  |  |  |  |  |
| Neonatal | 333 (77.8) | 146 (84.9) | 78 (79.6) | 57 (63.3) | 15 (55.6) | 18 (85.7) | 19 (95.0) | <0.001 |
| Other<sup>b</sup> | 95 (22.2) | 26 (15.1) | 20 (20.4) | 33 (36.7) | 12 (44.4) | 3 (14.3) | 1 (5.0) |  |
| **Gestational age (weeks)** | 33 [29 - 37] | 32 [29 - 36] | 30 [28 - 34] | 37 [32 - 37] | 36 [30 - 37] | 36 [32 - 39] | 36 [34 - 37] | <0.001 |
| **Underlying conditions** |  |  |  |  |  |  |  |  |
| Neonatal jaundice | 80 (18.7) | 29 (16.9) | 27 (27.6) | 13 (14.4) | 4 (14.8) | 5 (23.8) | 2 (10.0) | 0.177 |
| HMD<sup>c</sup>/ RDS<sup>d</sup> | 71 (16.6) | 23 (13.4) | 32 (32.7) | 9 (10.0) | 3 (11.1) | 2 (9.5) | 2 (10.0) | 0.001 |
| NEC<sup>e</sup> | 23 (5.4) | 15 (8.7) | 5 (5.1) | 2 (2.2) | 0 (0) | 1 (4.8) | 0 (0) | 0.204 |
| Cardiac abnormalities | 26 (6.1) | 12 (7.0) | 9 (9.2) | 3 (3.3) | 0 (0) | 0 (0) | 2 (10.0) | 0.258 |
| Other | 151 (35.3) | 67 (39.0) | 40 (40.8) | 24 (26.7) | 7 (33.3) | 7 (33.3) | 6 (30.0) | 0.261 |
| **Current weight<sup>f</sup>** |  |  |  |  |  |  |  |  |
| ≤1500 g | 100 (44.4) | 44 (45.4) | 38 (67.9) | 9 (23.1) | 3 (27.3) | 4 (33.3) | 2 (20.0) | <0.001 |
| 1501-2499 g | 52 (23.1) | 29 (29.9) | 10 (17.9) | 2 (5.1) | 3 (27.3) | 4 (33.3) | 4 (40.0) |  |
| ≥2500 g | 73 (32.4) | 24 (24.7) | 8 (14.3) | 28 (71.8) | 5 (45.5) | 4 (33.3) | 4 (40.0) |  |
| **Infection type** |  |  |  |  |  |  |  |  |
| Bloodstream | 425 (99.3) | 172 (100) | 97 (99) | 89 (98.9) | 26 (96.3) | 21 (100) | 20 (100) | 0.180 |
| Meningitis | 3 (0.7) | 0 (0) | 1 (1.0) | 1 (1.1) | 1 (3.7) | 0 (0) | 0 (0) |  |
| **Infection timing<sup>g</sup>** |  |  |  |  |  |  |  |  |
| Early-onset | 77 (18.2) | 24 (14.1) | 12 (12.2) | 15 (16.9) | 8 (30.8) | 8 (38.1) | 10 (50.0) | <0.001 |
| Late-onset | 347 (81.8) | 146 (85.9) | 86 (87.8) | 74 (83.1) | 18 (69.2) | 13 (61.9) | 10 (50.0) |  |
| **Healthcare-transmission rate<sup>h</sup>** |  |  |  |  |  |  |  |  |
| SNP-EPI method | 131 (30.6) | 72 (41.9) | 47 (48.0) | 10 (11.1) | 0 (0) | 0 (0) | 2 (10) | <0.001 |
| AST-EPI method | 77 (18.0) | 39 (22.7) | 28 (28.6) | 10 (11.1) | 0 (0) | 0 (0) | 0 (0) | <0.001 |
| **In-hospital outcome** |  |  |  |  |  |  |  |  |
| Alive<sup>i</sup> | 189 (68.2) | 74 (64.3) | 36 (50.7) | 46 (92.0) | 10 (71.4) | 14 (87.5) | 9 (81.8) | <0.001 |
| Died | 88 (31.8) | 41 (35.7) | 35 (49.3) | 4 (8.0) | 4 (28.6) | 2 (12.5) | 2 (18.2) |  |

a: IQR = interquartile range; b: includes any unit where infants <6 months were admitted; c: HMD = Hyaline membrane disease; d: RDS = respiratory distress syndrome; e: NEC = necrotising enterocolitis; f: weight at positive culture specimen collection; g: early onset = positive culture specimen collection from day of admission up to day 2 of life, late onset = specimen collection on or after day 3 of life; h: Rate calculated as the number of cases of infection allocated to a cluster by the specified method divided by all cases with both pathogen antimicrobial susceptibility and whole genome sequence data; i: Alive includes patients who were alive on discharge or transferred to another facility

Using the SNP-EPI criteria, we identified 39 clusters, with the majority caused by *K. pneumoniae* (46%, n=18), *A. baumannii* (38%, n=15) and *S. aureus* (13%, n=5), and a single cluster of *E. faecium*. The largest cluster was caused by NDM-1-producing ST152 *K. pneumoniae* strains in hospital C (n=15) alongside a smaller simultaneous cluster of OXA-1-producing ST307 *K. pneumoniae* (n=4) (**Fig 1, Fig S2**). Medium-sized *K. pneumoniae* clusters of OXA-1-producing ST307 (n=6) in hospital E, OXA-1/181-producing ST39 (n=6) in hospital B and OXA-1-producing ST405 (n=7) in hospital D were detected. Smaller *K. pneumoniae* clusters caused by ST348 and OXA-1/181-producing ST17 were recorded at different time points in hospital B. Overall, 11 of the 18 (61%) *K. pneumoniae* clusters had one or more isolates that were phenotypically resistant to carbapenems (**Table 2**). *A. baumannii* clusters were primarily caused by MDR OXA-23- and/or NDM-1-producing ST1 and ST2 strains (**Fig S3**), with notable clusters of ST2 in hospital B (n=7 and n=6) and ST1 in hospital E (n=7). Clusters of *S. aureus* and *E. faecium* (data not shown) were small (≤2 cases per cluster), with four of five *S. aureus* clusters caused by methicillin-resistant strains (**Fig S4**). No clusters of *E. faecalis* or *E. coli* were detected using the SNP-EPI criteria.

**Fig 1:**
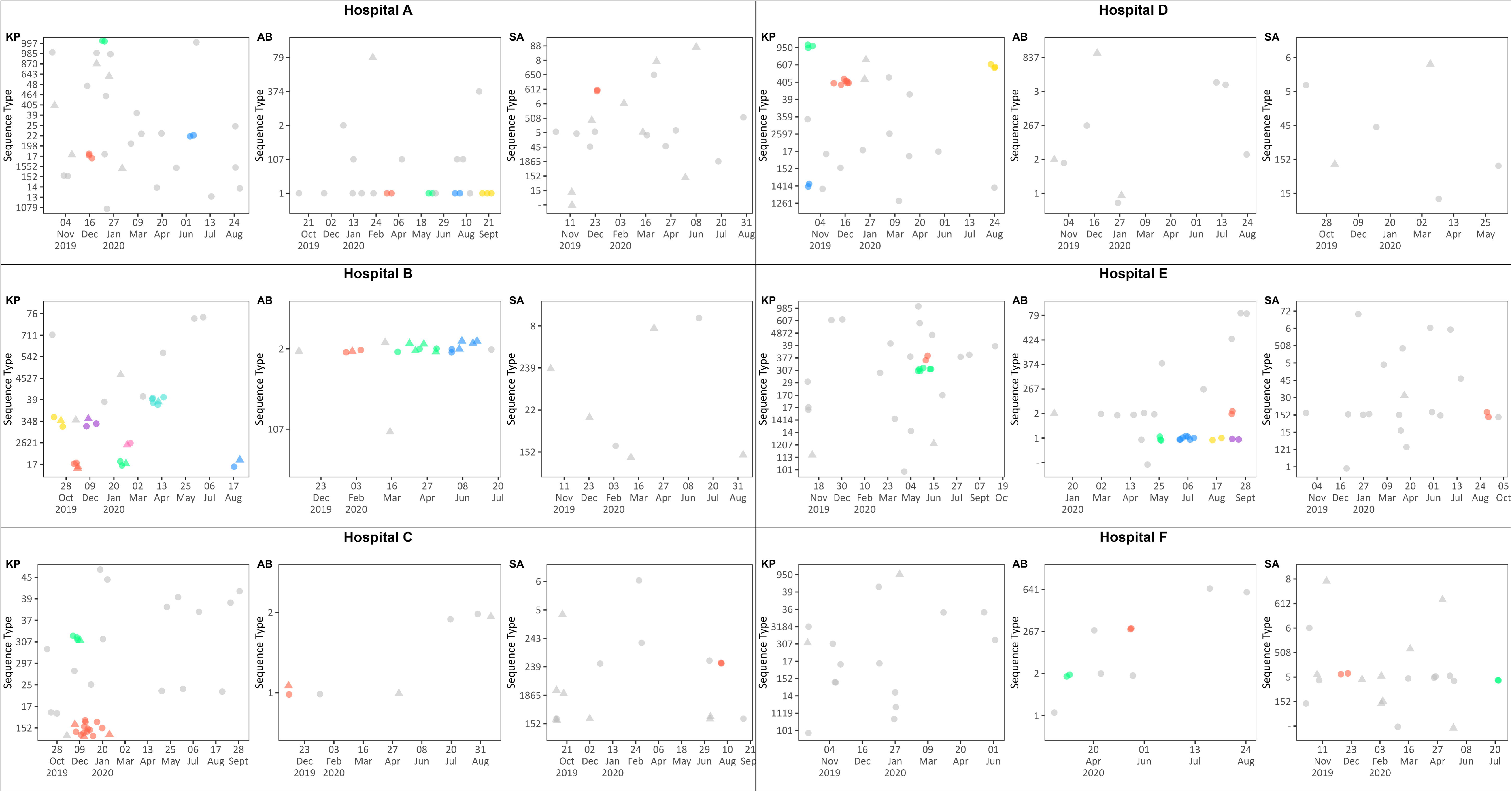
*K. pneumoniae*, *A. baumannii* and *S. aureus* infection clusters detected using the SNP-EPI cluster criteria for hospitals (A-F), by sequence type and time, September 2019 through to October 2020. Key: KP: *K. pneumoniae*; AB: *A. baumannii*; SA: *S. Aureus* Colors denote distinct clusters, circles and triangles denote case-patients in neonatal and other wards, respectively. Other wards included general pediatric wards, pediatric intensive care units (ICU), general ICU and/or high care wards. The grey dots (bg) represent cases that were not categorized as part of clusters by the SNP-EPI criteria.

**Table 2:**
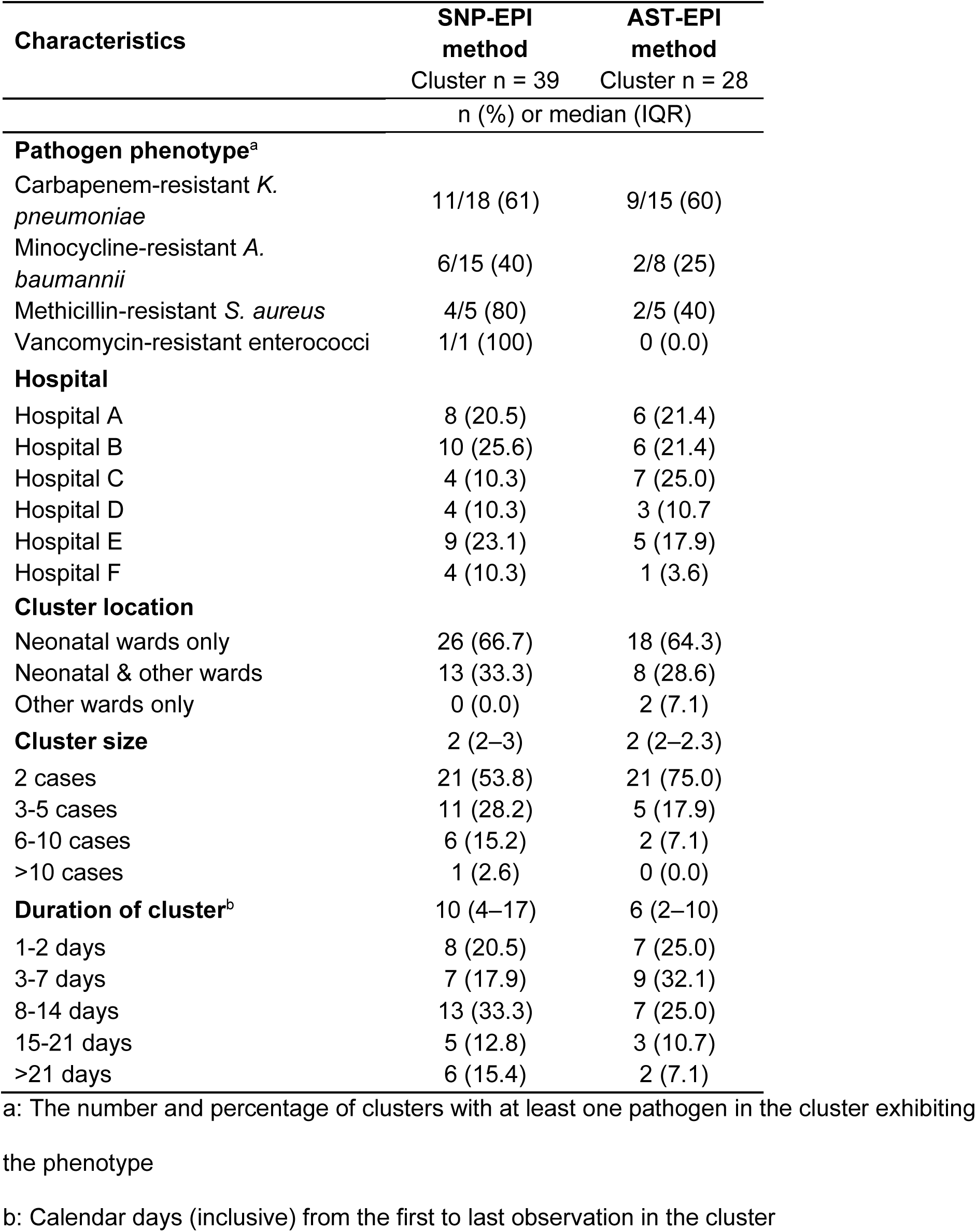
Characteristics of clusters at six lower-tier hospitals in South Africa, September 2019 - October 2020.

In the univariable logistic regression analysis, older infants at specimen collection (OR 0.98, 95% CI 0.96-0.99, p=0.002) and those with a weight of ≥2500 g at specimen collection (OR 0.20, 95% CI 0.09-0.44, p<0.001) were less likely to be in a cluster, while those with an umbilical line were more likely to be in a cluster (OR 2.02, 95% CI 1.03-3.90) (**Table S2**). In the multivariable model, infants with a weight of ≥2500 g were 80% less likely to be in a cluster (adjusted OR 0.20, 95% CI 0.04-0.83, p=0.033).

The overall healthcare transmission rate using the primary SNP-EPI criteria was 31% (131/428 cases) (**Table 1**); this was highest for *A. baumannii* (48%, 47/98) followed by *K. pneumoniae* (42%, 72/172), *S. aureus* (11%, 10/90) and *E. faecium* (10%, 2/10) (p<0.001). For each pathogen, the healthcare transmission rate varied by facility (range, 5%-95%) (**Fig 2**). Relaxing the SNP-EPI criteria resulted, on average, in the inclusion of 1-2 additional cases within clusters, merging of clusters and/or detection of new clusters (**Fig S5-S7**), which increased the estimated healthcare transmission rate (**Fig 2**). Cluster sizes and/or patterns changed more when increasing the threshold duration between specimen collection dates than when increasing SNP cut-offs (**Fig 2, Fig S5-S7**). Clusters of *E. faecalis* or *E. coli* were not detected when varying the SNP-EPI criteria.

**Fig 2:**
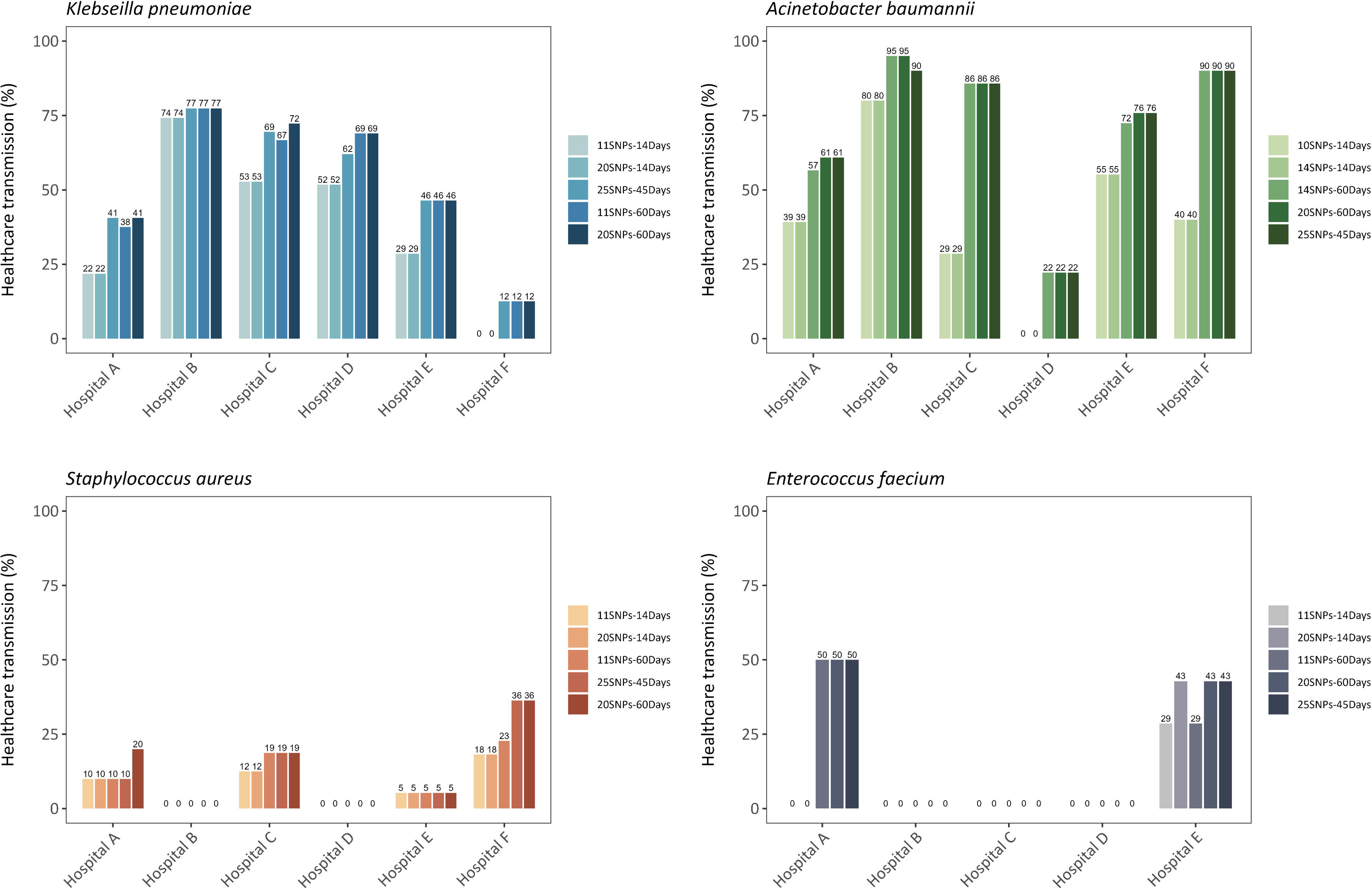
Healthcare transmission rates (i.e. proportion of cases linked to a cluster) by pathogen and hospital when varying SNP-EPI parameters.

We detected 28 AST-EPI clusters compared to 39 SNP-EPI clusters (**Table 2**). Most of the clusters that were missed when applying the AST-EPI criteria were caused by *A. baumannii* (15 versus 8). The AST-EPI criteria resulted in fragmented clusters or entirely different clusters (**Fig 3**). Overall, clusters identified using the SNP-EPI criteria were larger. The proportion of clusters with the smallest size (i.e., 2 cases per cluster) was higher when applying AST-EPI criteria (75%, 21/28) than by applying SNP-EPI criteria (53.8%, 21/39).

**Fig 3:**
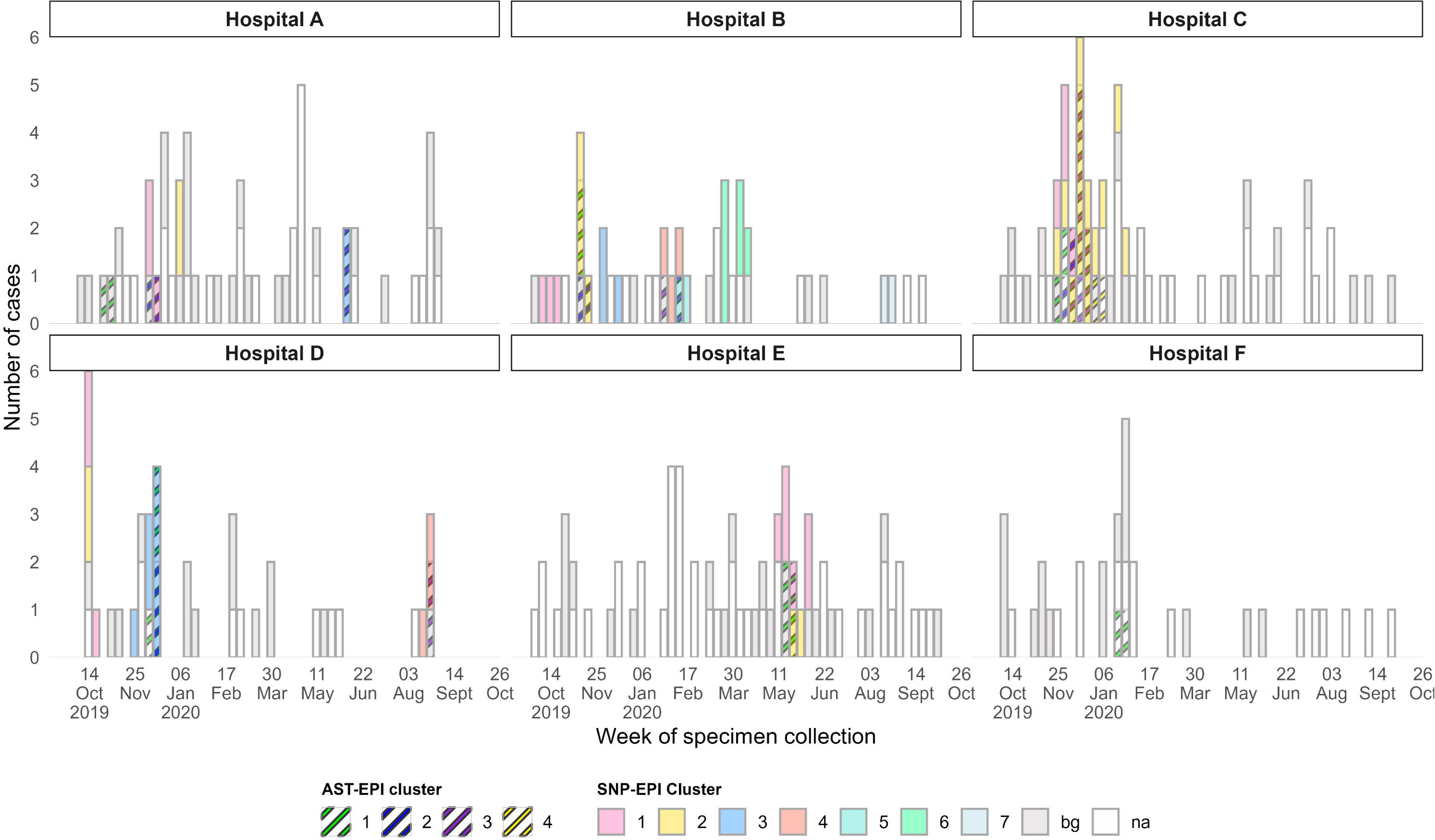
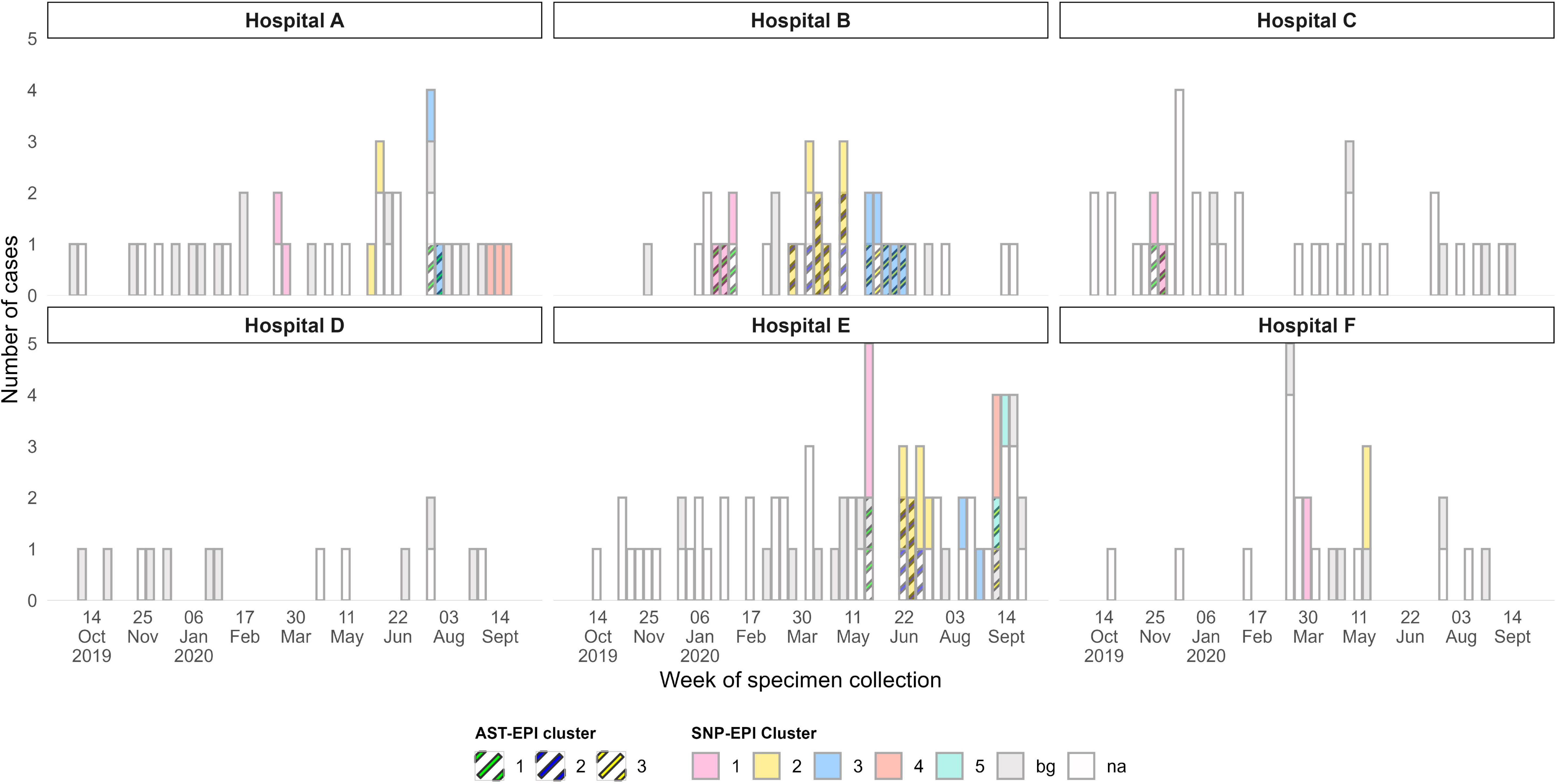
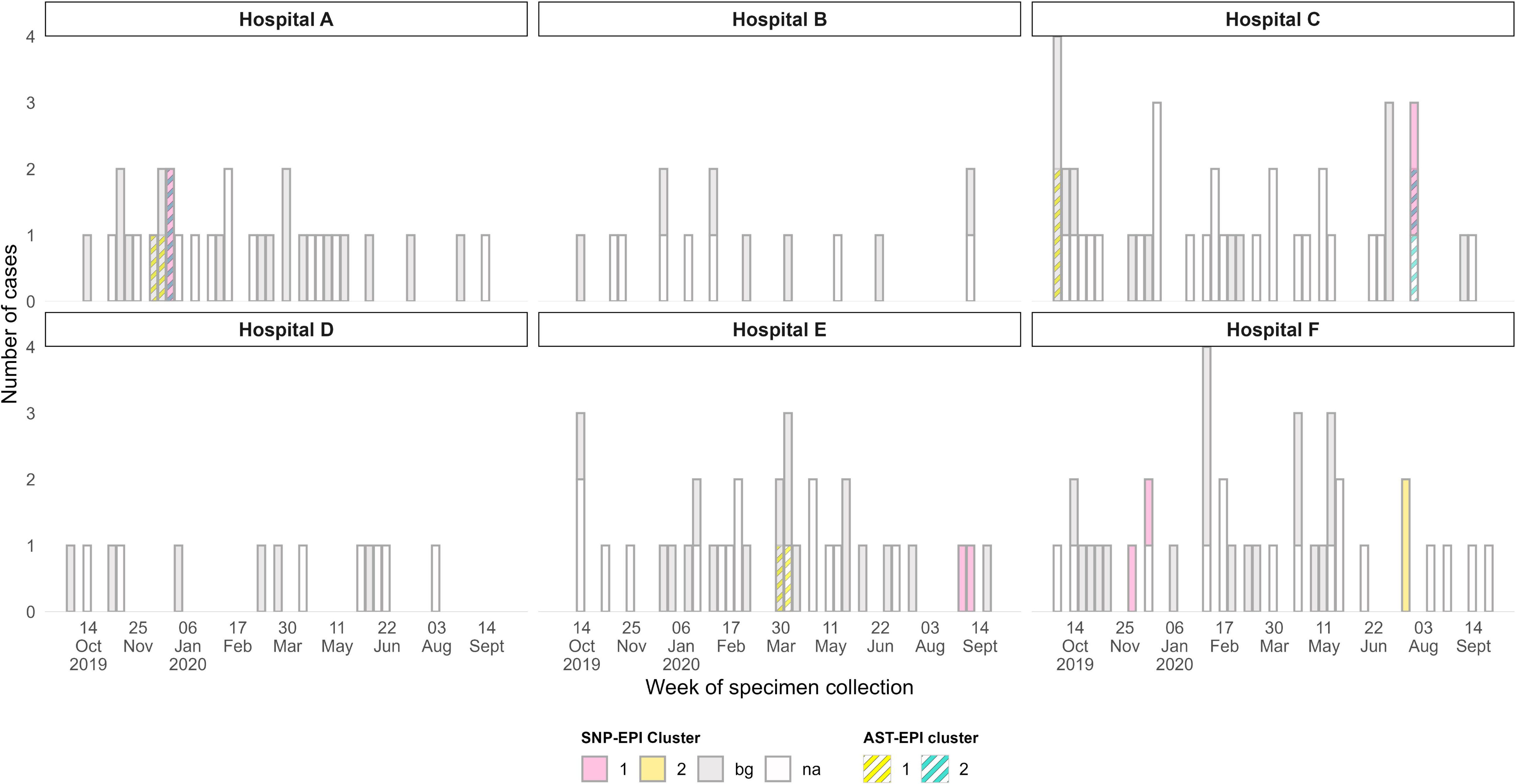
Infection clusters of A) *K. pneumoniae*, B) *A. baumannii*, and C) *S. aureus* detected in Hospital A-F using the SNP-EPI and AST-EPI cluster criteria, September 2019-October 2020. Boxes shaded in grey (bg) represent cases with both pathogen antimicrobial susceptibility testing (AST) and whole genome sequence (WGS) data but were not linked to clusters. White boxes (na) represent case-patients without AST and/or WGS data and thus excluded from the analysis

SNP-EPI clusters also occurred over a longer period than AST-EPI clusters (median, 10 days [IQR: 4-17] versus 6 days [IQR: 2-10]). The healthcare transmission rate was lower when applying the AST-EPI criteria overall (18% versus 31%), to *K. pneumoniae* (23% versus 42%) and to *A. baumannii* (29% versus 48%), but the rate remained the same for *S. aureus* (i.e. 11%).

## Discussion

The national incidence of culture-confirmed neonatal infections in South Africa was previously documented to be high, with a substantial proportion caused by Gram-negative bacteria [2]. Although it has been speculated that transmission events mediated by healthcare workers and/or environmental contamination play a major role, our study aimed to provide robust evidence for this by combining genomic and epidemiological data. We found that almost a third of the infection episodes analyzed in our study probably occurred through healthcare transmission. This was similar to the 31% reported in an Australian study conducted among in-patients infected with MDR pathogens, including methicillin-resistant *S. aureus*, ceftriaxone-resistant *E. coli* and *K. pneumoniae*, and vancomycin-resistant enterococci [4]. In a systematic review of studies that retrospectively identified clusters using genomic and epidemiological data, the average putative healthcare transmission was 23% overall, with 0% in some studies and rates as high as 61.6% in others [14], demonstrating that transmission patterns probably differ by pathogen and setting. While results from these studies provide some insight into the relative contribution of healthcare transmission to the burden of disease, we acknowledge that direct comparisons are inherently limited by methodological differences.

Compared to other pathogens, we found substantially higher healthcare transmission of *K. pneumoniae* (up to 74%) or *A. baumannii* (up to 80%) at some facilities. These findings are consistent with the predominance of Gram-negative infections among neonates in LMICs, including South Africa [2, 17]. Prematurity and low birth weight play a substantial role in the vulnerability of newborns to Gram-negative bacterial infections. We found that infants who weighed ≥2500 g were 80% less likely to be in a cluster. A combination of factors, such as immunological immaturity, impaired physical barriers and healthcare-related risk factors such as invasive procedures which occur during prolonged hospital stays, create multiple entry points and opportunities for invasive infection particularly in low-weight premature infants [18–20]. Pathogen-specific factors also contribute. *K. pneumoniae* and *A. baumannii* are frequently multidrug resistant and can outcompete susceptible bacteria under empirical broad-spectrum antibiotic pressure, which is common in neonatal care. Furthermore, both species are highly adapted to persist in hospital environments and colonize hosts for prolonged periods, establishing a constant reservoir for infection and outbreaks, especially where IPC measures are suboptimal [21–23]. The high transmission rates of these MDR pathogens places a significant burden on patients and the healthcare system due to treatment challenges and high mortality risk. Consistent with this, infants in our study infected with *A. baumannii* and *K. pneumoniae* had a higher crude in-hospital mortality compared to infants with other pathogens. In contrast, we found that clusters of *S. aureus* and *E. faecium* were generally small, and no clusters of *E. faecalis* or *E. coli* were detected. Infections caused by these latter species may be more sporadic in nature and healthcare transmission may therefore play a comparatively minor role in their overall burden.

A high proportion of clusters detected in our study were caused by MDR strains. The largest was a prolonged polyclonal outbreak caused by NDM-1-producing ST152 *K. pneumoniae* and OXA-181 ST307 *K. pneumoniae*, which was detected during the Baby GERMS-SA surveillance study and has been described previously [24]. We also found other notable clusters caused by high-risk, globally distributed, carbapenem-resistant *K. pneumoniae* clones, including ST307, ST39, ST405, ST348, and ST17. These clones often possess various additional antimicrobial resistance (AMR) determinants and are frequently associated with outbreaks worldwide [25, 26]. IMP-38-producing ST307, NDM-1-producing ST17, and NDM-producing ST348 outbreaks have been documented in neonatal units in other regions [27–29]. The OXA-181 producing ST307 strain, predicted to have been introduced into private-sector hospitals in South Africa in 2023, rapidly spread via intra- and inter-hospital transmission and is now considered endemic in South Africa [30, 31]. While ST405 is also recognized globally as a MDR lineage, there are fewer documented outbreaks compared to ST307, and none reported in neonatal units [32]. Similarly, *A. baumannii* clusters were caused by AMR-associated clones, specifically the globally dominant ST1 and ST2 lineages [33]. While outbreaks of MDR *A. baumannii* in neonatal units are well-documented, the genomic characteristics of the strains involved are less frequently reported. Nonetheless, regional neonatal outbreaks of MDR OXA-23- and NDM-1-producing ST1 outbreaks have been reported in Botswana and the KwaZulu-Natal Province of South Africa [34, 35]. Our results confirm the presence of high-risk clones that are spread by healthcare transmission in South African neonatal units. The presence of these clones and the outbreaks they cause present a major public health problem, particularly in the lower-tier hospitals included in this study. These facilities currently lack the resources for routine genomic surveillance needed to detect high-risk lineage clusters, which in turn elevates the risk of mortality due to limited treatment options for these MDR organisms. In this study, infants with *A. baumannii* and *K. pneumoniae* infections had much higher risk of in-hospital mortality than those with *S. aureus* and other pathogens.

Previous studies that considered within-host diversity, pathogen mutation rates, and/or epidemiological information proposed SNP cutoffs for recent transmission, for example ≤25 whole-genome SNPs for MRSA and ≤11 SNPs for *K. pneumoniae* [36, 37]. Despite this, a wide range of SNP cutoffs continue to be used in outbreak investigations [14]. Due to the absence of a standardized definition for outbreaks, we examined the impact of adjusting both the SNP cutoff (to define isolate relatedness) and the time interval between specimen collection dates. Modifying these parameters changed the clustering patterns and increased the estimated rates of healthcare transmission, with the duration between specimen collection dates having a more significant influence. This indicates that while our primary definition is probably suitable for rapid-onset outbreaks, which are common in our setting, it may overlook prolonged outbreaks associated with contaminated environmental reservoirs or those involving pathogens with a faster genetic evolutionary rate [3, 34, 38]. Failing to recognize ongoing transmission could result in inadequate IPC measures being implemented, leading to higher infection rates. Nevertheless, our primary definition successfully identified the majority of clusters compared to alternative criteria, although it split some clusters and excluded certain individuals from transmission chains. To facilitate cluster detection, accurate measurement of healthcare transmission, valid external comparisons, and monitoring of temporal changes, there is a need for the development and adoption of standardized, evidence-based, and universally applicable methodologies for clinically significant pathogens.

In our analysis, we found that all but two concurrently occurring clusters in a single facility were not recognized during the surveillance period. This highlights the inadequacies of relying solely on healthcare workers’ perceptions or routine surveillance data, particularly in settings with high infection incidence. We directly compared conventional AST-based clusters with those defined by WGS SNP data. Consistent with prior studies, we discovered that AST patterns missed several SNP-defined clusters and/or fragmented them into multiple smaller clusters [39]. This highlights the limited discriminatory power of phenotypic pathogen profiles, which may misinterpret clonal spread due to convergent resistance patterns, horizontal gene transfer, or differential expression of resistance genes. In contrast, WGS provided a higher-resolution view of the isolates, revealing larger, more persistent clusters and enabling differentiation of concurrent outbreaks caused by distinct sequence types within the same hospital. Despite WGS being considered the gold standard, the implementation of genomic surveillance to inform IPC and/or diagnostic and antimicrobial stewardship is still challenged by long turnaround times, high costs, and the need for specialized bioinformatics expertise [40]. Measures to address these challenges are actively being explored, including the possible adoption of rapid technologies such nanopore sequencing and the development of culture-independent metagenomics [41, 42]. Regional genomic hubs have been proposed to reduce cost, improve resource efficiency, and provide technical training on analysis and interpretation of WGS data to local laboratories and/or facilities [40]. Although AST is relatively inexpensive and remains crucial as a complement to WGS for phenotypic resistance confirmation and clinical treatment guidance, its utility for definitive outbreak delineation is limited. Nonetheless, AST data can still be a valuable complement to WGS, particularly in detection of emerging strains with novel resistance profiles and unknown genetic determinants [43].

Our study is subject to several limitations that impact on the precision and generalizability of our findings on healthcare transmission. First, the analysis was limited by isolate availability, as nearly two-thirds (58%) of isolates from case-patients were unavailable for WGS. This substantial missing data introduced a sampling bias. Depending on whether the excluded infections belonged to a cluster or were isolated cases, this bias could potentially result in an overestimation or underestimation of the calculated transmission proportions, potentially obscuring the full reality of transmission patterns across the hospitals. Secondly, we lacked crucial hospital utilization data (such as patient-days) to contextualize our findings, precluding the calculation of adjusted transmission rates and hindering direct comparison with other studies. Furthermore, the scope of the study was restricted to only invasive infections (bloodstream and central nervous system) caused by the most common ESKAPE pathogens. Our estimates may not fully reflect the total burden of healthcare transmission, as we did not include colonization screening, less severe infections, or infections caused by other clinically-relevant bacterial and fungal pathogens. Finally, the retrospective design limited our capacity to integrate granular ward-level epidemiological or environmental data, which would have been essential for identifying specific infection sources or drivers of infection acquisition. Although we found that infants with a higher body weight were less likely to be in a cluster, our study was not specifically powered for this analysis, and thus we cannot rule out or rule in any of the patient- or healthcare-level factors in their role in facilitating healthcare transmission or outbreaks. While this design prevents direct linkage of genomic findings to immediate IPC intervention outcomes, the genomic resolution achieved provides a baseline for prospective studies and highlights the scale of the problem, suggesting on average around one-third of infections were associated with nosocomial transmission.

In conclusion, our study provides compelling evidence that healthcare-associated transmission contributes substantially to the burden of invasive bacterial infections among infants in South Africa, particularly driven by MDR *K. pneumoniae* and *A. baumannii*. WGS offers substantially higher resolution than conventional AST, effectively delineating complex transmission chains and simultaneously identifying crucial resistance determinants. Genomic approaches should be integrated into clinical and public health practice for robust outbreak detection and response, specifically as an outbreak investigative response to signals such as a rise in case numbers, high rates of specific MDR pathogens, or the detection of new resistance patterns. While ongoing genomic surveillance (i.e. sequencing most isolates) may be the goal, an investigative strategy is a pragmatic and impactful starting point for low-resource settings. Strengthening IPC practices remains an essential core action, but WGS complements these efforts by pinpointing where and when interventions are most needed, enabling timely, evidence-based action to limit the spread of pathogens and improve outcomes for vulnerable newborns in low-resource healthcare settings.

## Data Availability

All data produced in the present study are available upon reasonable request to the authors.

## Acknowledgements

We thank the members of Baby GERMS-SA: Dora Nginza Hospital: Phunyezwa Mzayiya, Shareef Abrahams, Vanessa Pearce, Zikhona Gabazana, Melissa Ngubane, Badikazi Matiwana; Klerksdorp Hospital: Omphile Mekgoe, Sebabatso Khantsi, Bernard Motsetse, Louisa Phalatse; Mankweng Hospital: Ruth Lekalakala, Tebogo Modiba, Molly Morapeli; National Institute for Communicable Diseases: Linda Erasmus, Danie Erwee, Juliet Paxton, Siyanda Dlamini, Marshagne Smith, Ruth Mpembe, Ntombi Dube, Relebohile Ramatsa, Thembekile Zwane, Sibongile Walaza, Erika van Schalkwyk; Queen Nandi Hospital: Constance Kapongo, Meluleki Mthimkhulu, Sandra Maphumulo, Dianette Pearce; Rob Ferreira Hospital: Lerato Motjale, Thulisile Maphosa, Greta Hoyland, Sindile Ntuli, Lesley Ingle; Tembisa Hospital: Harishia Naidoo, Ramatlhwa Kekana, Dina Pombo. We are grateful to the National Institute for Communicable Diseases Sequencing Core Facility.

## Funding statement

This publication is based on research funded in part by the Gates Foundation (INV-047481). The findings and conclusions contained within are those of the authors and do not necessarily reflect positions or policies of the Gates Foundation.

## Author contributions

**Conceptualization:** LS, HI, RM1, KEH, NPG; **Data Curation:** RM2, SK, OP, STM, VCQ; **Formal analysis:** LS, HI, RM1, SK**; Visualization:** LS, SK; **Funding acquisition:** LS, HI, RM1, NPG; **Supervision:** NPG; **Writing – original draft preparation:** LS; Writing – review & editing: LS, HI, RM1, RM2, SK, KEH, OP, STM, VCQ, NPG

## Notes

### Competing Interest Statement

The authors have declared no competing interest.

## References

[1] Mahtab S, Madhi SA, Baillie VL, et al. Causes of death identified in neonates enrolled through Child Health and Mortality Prevention Surveillance (CHAMPS), December 2016 –December 2021. PLOS Global Public Health 2023; 3: e0001612.

[2] Mashau RC, Meiring ST, Dramowski A, et al. Culture-confirmed neonatal bloodstream infections and meningitis in South Africa, 2014-19: a cross-sectional study. Lancet Glob Health 2022; 10: e1170–e1178.

[3] Quan KA, Sater MRA, Uy C, et al. Epidemiology and genomics of a slow outbreak of methicillin-resistant *Staphyloccus aureus* (MRSA) in a neonatal intensive care unit: Successful chronic decolonization of MRSA-positive healthcare personnel. Infect Control Hosp Epidemiol 2023; 44: 589–596.

[4] Sherry NL, Gorrie CL, Kwong JC, et al. Multi-site implementation of whole genome sequencing for hospital infection control: A prospective genomic epidemiological analysis. Lancet Reg Health West Pac 2022; 23: 100446.

[5] Sherry NL, Gorrie CL, Kwong JC, et al. Pilot study of a combined genomic and epidemiologic surveillance program for hospital-acquired multidrug-resistant pathogens across multiple hospital networks in Australia. Infect Control Hosp Epidemiol 2021; 42: 573–581.

[6] Sundermann AJ, Kumar P, Griffith MP, et al. Real-Time Genomic Surveillance for Enhanced Healthcare Outbreak Detection and Control: Clinical and Economic Impact. Clinical Infectious Diseases. DOI: 10.1093/cid/ciaf216.

[7] Meiring S, Quan V, Mashau R, et al. Pathogen aetiology and risk factors for death among neonates with bloodstream infections at lower-tier South African hospitals: a cross-sectional study. Lancet Microbe 2025; 6: 100989.

[8] Kwenda S, Allam M, Khumalo Z.T.H, et al. Jekesa: an automated easy-to-use pipeline for bacterial whole genome typing. github, https://github.com/stanikae/jekesa (2020, accessed 4 November 2025).

[9] Feldgarden M, Brover V, Gonzalez-Escalona N, et al. AMRFinderPlus and the Reference Gene Catalog facilitate examination of the genomic links among antimicrobial resistance, stress response, and virulence. Sci Rep 2021; 11: 12728.

[10] Bortolaia V, Kaas RS, Ruppe E, et al. ResFinder 4.0 for predictions of phenotypes from genotypes. Journal of Antimicrobial Chemotherapy 2020; 75: 3491–3500.

[11] Mendes I, Griffiths E, Manuele A, et al. hAMRonization: Enhancing antimicrobial resistance prediction using the PHA4GE AMR detection specification and tooling. bioRxiv. Epub ahead of print 11 March 2024. DOI: 10.1101/2024.03.07.583950.

[12] Seemann Torsten. scapper: Whole genome core alignments from multiple draft genomes. github, https://github.com/tseemann/scapper (2014, accessed 4 November 2025).

[13] Kwenda S, Shuping L, Mashau R, Ismail H, Govender N.P. SNP2Cluster: A core SNP and K-means clustering-based tool for enhanced transmission cluster detection in outbreak scenarios. Zenodo 2024; DOI: 10.5281/zenodo.13976252.

[14] Sundermann AJ, Chen J, Miller JK, et al. Whole-genome sequencing surveillance and machine learning for healthcare outbreak detection and investigation: A systematic review and summary. Antimicrobial Stewardship & Healthcare Epidemiology 2022; 2: e91.

[15] Spencer MD, Winglee K, Passaretti C, et al. Whole Genome Sequencing detects Inter-Facility Transmission of Carbapenem-resistant *Klebsiella pneumoniae*. Journal of Infection 2019; 78: 187–199.

[16] Thoma R, Seneghini M, Seiffert SN, et al. The challenge of preventing and containing outbreaks of multidrug-resistant organisms and *Candida auris* during the coronavirus disease 2019 pandemic: report of a carbapenem-resistant *Acinetobacter baumannii* outbreak and a systematic review of the literature. Antimicrob Resist Infect Control 2022; 11: 12.

[17] Hallmaier-Wacker LK, Andrews A, Nsonwu O, et al. Incidence and aetiology of infant Gram-negative bacteraemia and meningitis: systematic review and meta-analysis. Arch Dis Child 2022; 107: 988–994.

[18] Shah BA, Padbury JF. Neonatal sepsis: an old problem with new insights. Virulence 2014; 5: 170–178.

[19] Flannery DD, Chiotos K, Gerber JS, et al. Neonatal multidrug-resistant gram-negative infection: epidemiology, mechanisms of resistance, and management. Pediatr Res 2022; 91: 380–391.

[20] Graham PL, Begg MD, Larson E, et al. Risk Factors for Late Onset Gram-Negative Sepsis in Low Birth Weight Infants Hospitalized in the Neonatal Intensive Care Unit. Pediatric Infectious Disease Journal 2006; 25: 113–117.

[21] Sultan AM, Seliem WA. Identifying Risk Factors for Healthcare-Associated Infections Caused by Carbapenem-Resistant *Acinetobacter baumannii* in a Neonatal Intensive Care Unit. Sultan Qaboos Univ Med J 2018; 18: e75–80.

[22] Sharma S, Das A, Banerjee T, et al. Adaptations of carbapenem resistant *Acinetobacter baumannii* (CRAB) in the hospital environment causing sustained outbreak. J Med Microbiol; 70. DOI: 10.1099/jmm.0.001345.

[23] Cai S, Wang Z, Han X, et al. The correlation between intestinal colonization and infection of carbapenem-resistant *Klebsiella pneumoniae*: A systematic review. J Glob Antimicrob Resist 2024; 38: 187–193.

[24] Magobo RE, Ismail H, Lowe M, et al. Outbreak of NDM-1– and OXA-181–Producing *Klebsiella pneumoniae* Bloodstream Infections in a Neonatal Unit, South Africa. Emerg Infect Dis; 29. DOI: 10.3201/eid2908.230484.

[25] Peirano G, Chen L, Kreiswirth BN, et al. Emerging Antimicrobial-Resistant High-Risk *Klebsiella pneumoniae* Clones ST307 and ST147. Antimicrob Agents Chemother; 64. DOI: 10.1128/AAC.01148-20.

[26] Karampatakis T, Zarras C, Pappa S, et al. Emergence of ST39 carbapenem-resistant *Klebsiella pneumoniae* producing VIM-1 and KPC-2. Microb Pathog 2022; 162: 105373.

[27] Strysko J, Thela T, Thubuka J, et al. Hyperendemic carbapenem-resistant Acinetobacter baumannii at a hospital in Botswana: Insights from whole-genome sequencing. Antimicrobial Stewardship & Healthcare Epidemiology 2023; 3: s115–s116.

[28] Jin Y, Shao C, Li J, et al. Outbreak of Multidrug Resistant NDM-1-Producing *Klebsiella pneumoniae* from a Neonatal Unit in Shandong Province, China. PLoS One 2015; 10: e0119571.

[29] Carlos CC, Masim MAL, Lagrada ML, et al. Genome Sequencing Identifies Previously Unrecognized *Klebsiella pneumoniae* Outbreaks in Neonatal Intensive Care Units in the Philippines. Clinical Infectious Diseases 2021; 73: S316–S324.

[30] Lowe M, Kock MM, Coetzee J, et al. *Klebsiella pneumoniae* ST307 with blaOXA-181, South Africa, 2014–2016. Emerg Infect Dis 2019; 25: 739–747.

[31] Salvador-Oke KT, Pitout JDD, Peirano G, et al. *Klebsiella pneumoniae* with carbapenemases: high prevalence of sequence type 307 with blaOXA181 in South African community hospitals. European Journal of Clinical Microbiology & Infectious Diseases 2024; 43: 2239–2244.

[32] Pano-Pardo JR, Ruiz-Carrascoso G, Navarro-San Francisco C, et al. Infections caused by OXA-48-producing *Klebsiella pneumoniae* in a tertiary hospital in Spain in the setting of a prolonged, hospital-wide outbreak. Journal of Antimicrobial Chemotherapy 2013; 68: 89–96.

[33] Shelenkov A, Akimkin V, Mikhaylova Y. International Clones of High Risk of *Acinetobacter Baumannii*—Definitions, History, Properties and Perspectives. Microorganisms 2023; 11: 2115.

[34] Strysko J, Thela T, Thubuka J, et al. Hyperendemic carbapenem-resistant *Acinetobacter baumannii* at a hospital in Botswana: Insights from whole-genome sequencing. Antimicrobial Stewardship & Healthcare Epidemiology 2023; 3: s115–s116.

[35] National Institute for Communicable Diseases. A cluster of multidrug-resistant *Acinetobacter baumannii* infections in a neonatal intensive care unit at a regional hospital in KwaZulu-Natal, August 2022. Johannesburg, https://www.nicd.ac.za/wp-content/uploads/2022/09/NICD-Monthly-Communiqué-Sept-2022.pdf (September 2022, accessed 7 October 2025).

[36] Coll F, Raven KE, Knight GM, et al. Definition of a genetic relatedness cutoff to exclude recent transmission of meticillin-resistant *Staphylococcus aureus*: a genomic epidemiology analysis. Lancet Microbe 2020; 1: e328–e335.

[37] Spencer MD, Winglee K, Passaretti C, et al. Whole Genome Sequencing detects Inter-Facility Transmission of Carbapenem-resistant *Klebsiella pneumoniae*. Journal of Infection 2019; 78: 187–199.

[38] Köser CU, Holden MTG, Ellington MJ, et al. Rapid Whole-Genome Sequencing for Investigation of a Neonatal MRSA Outbreak. New England Journal of Medicine 2012; 366: 2267–2275.

[39] Harris SR, Cartwright EJ, Török ME, et al. Whole-genome sequencing for analysis of an outbreak of meticillin-resistant *Staphylococcus aureus*: a descriptive study. Lancet Infect Dis 2013; 13: 130–136.

[40] Jauneikaite E, Baker KS, Nunn JG, et al. Genomics for antimicrobial resistance surveillance to support infection prevention and control in health-care facilities. Lancet Microbe 2023; 4: e1040–e1046.

[41] Charalampous T, Alcolea-Medina A, Snell LB, et al. Evaluating the potential for respiratory metagenomics to improve treatment of secondary infection and detection of nosocomial transmission on expanded COVID-19 intensive care units. Genome Med 2021; 13: 182.

[42] Loman NJ, Constantinidou C, Christner M, et al. A Culture-Independent Sequence-Based Metagenomics Approach to the Investigation of an Outbreak of Shiga-Toxigenic *Escherichia coli* O104:H4. JAMA 2013; 309: 1502.

[43] Köser CU, Ellington MJ, Cartwright EJP, et al. Routine Use of Microbial Whole Genome Sequencing in Diagnostic and Public Health Microbiology. PLoS Pathog 2012; 8: e1002824.

