## supplementary_information for "Enhanced detection of neonatal invasive infection clusters in South Africa using epidemiological and genomic surveillance data"

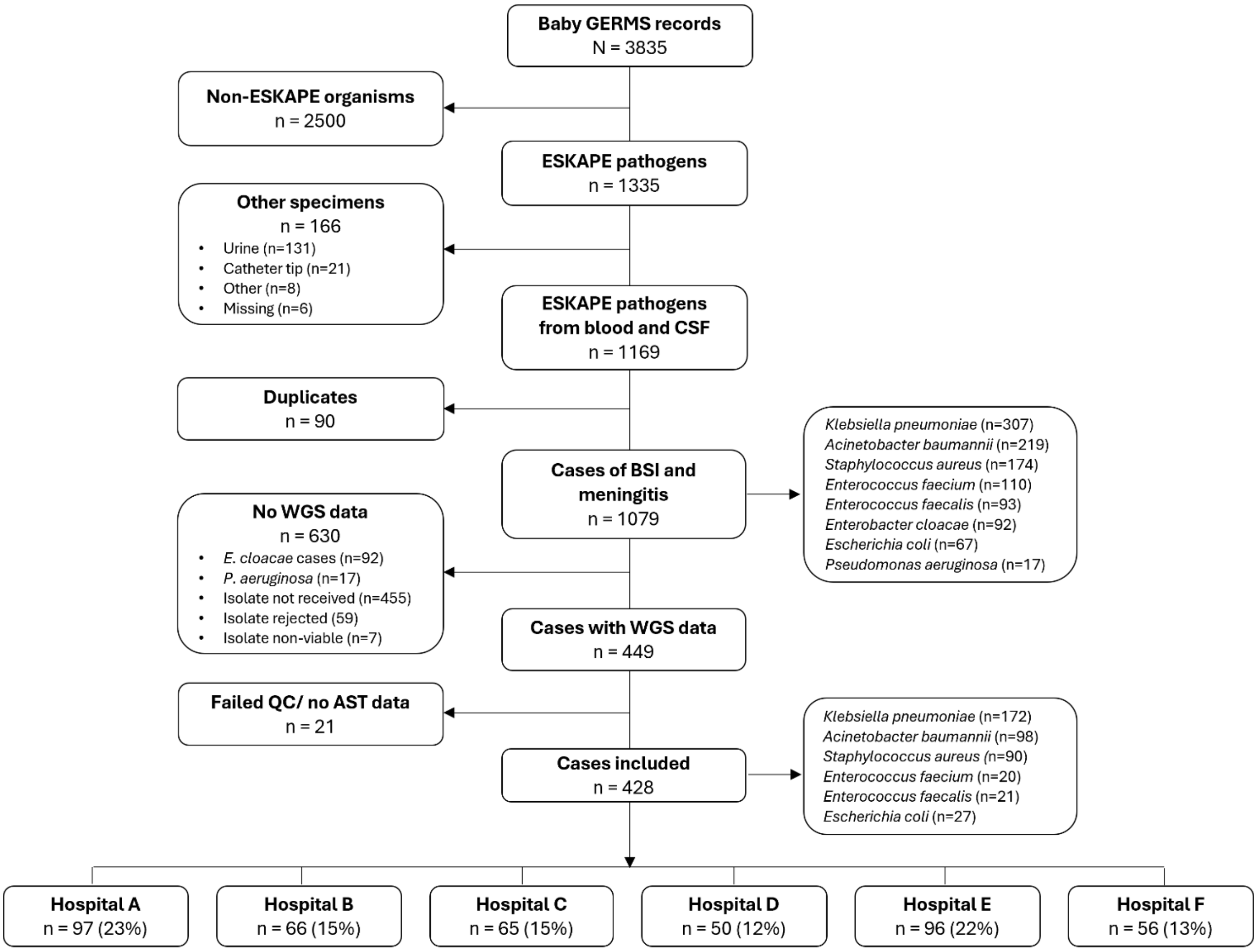


**Fig S1:** Cases of BSI and meningitis at six Baby GERMS-SA sentinel hospitals included in the study

Key; ESKAPE = *Enterococcus faecium* and *faecalis*, *Staphylococcus aureus*, *Klebsiella pneumoniae*, *Acinetobacter baumannii*, *Pseudomonas aeruginosa*, *Enterobacter cloacae* and *Escherichia coli*; CSF = Cerebrospinal fluid; BSI = bloodstream infection; WGS = Whole genome sequencing; QC = Quality control; AST = Antimicrobial susceptibility testing

**Table S1:** Odds of death by pathogen among patients with bloodstream infection or meningitis at six sentinel hospitals in South Africa, September 2019–October 2020, n=428

| Pathogen | n | OR[a] (95% CI[b]) | p-value |
|---|---|---|---|
| *Staphylococcus aureus* | 90 | Ref | |
| *Klebsiella pneumoniae* | 172 | 6.37 (2.14-18.96) | 0.001 |
| *Acinetobacter baumannii* | 98 | 11.18 (3.63-34.35) | <0.001 |
| *Escherichia coli* | 27 | 4.6 (0.98-21.57) | 0.053 |
| *Enterococcus faecalis* | 21 | 1.64 (0.27-9.93) | 0.589 |
| *Enterococcus aecium* | 20 | 2.55 (0.41-16.11) | 0.405 |

a: OR = odds ratio; b: CI = confidence interval

**Table S2:** Logistic regression model of factors associated with being in a SNP-EPI cluster at six sentinel hospitals in South Africa, September 2019–October 2020, n=428

| | Univariable | | | Multivariable | | |
|---|---|---|---|---|---|---|
| Characteristic | n | OR[a] (95% CI[b]) | p-value | n | aOR[c] 95% CI | p-value |
| **Age (days)** | 424 | 0.98 (0.96-0.99) | 0.002 | 196 | 0.98 (0.94-1.01) | 0.300 |
| **Sex** | | | | | | |
| Female | 191 | Ref | | 90 | Ref | |
| Male | 221 | 0.85 (0.56-1.30) | 0.400 | 106 | 1.1 (0.56-2.15) | 0.800 |
| **HIV exposed** | | | | | | |
| No | 148 | Ref | | 124 | Ref | |
| Yes | 84 | 0.86 (0.48-1.52) | 0.600 | 72 | 0.62 (0.30-1.26) | 0.200 |
| **Gestation age (weeks)** | | | | | | |
| ≤27 | 28 | Ref | | 24 | Ref | |
| 28-31 | 70 | 0.89 (0.37-2.19) | 0.800 | 56 | 0.68 (0.24-1.91) | 0.500 |
| 32-36 | 70 | 0.74 (0.30-1.83) | 0.500 | 60 | 1.05 (0.32-3.44) | >0.900 |
| ≥37 | 68 | 0.23 (0.08-0.62) | 0.004 | 56 | 1.71 (0.13-4.10) | 0.700 |
| **Weight (grams)**[d] | | | | | | |
| ≤1500 | 100 | Ref | | 85 | Ref | |
| 1501-2499 | 52 | 1.14 (0.58-2.25) | 0.700 | 47 | 0.91 (0.37-2.24) | 0.800 |
| ≥2500 | 73 | 0.20 (0.09-0.44) | <0.001 | 64 | 0.20 (0.04-0.83) | 0.033 |
| **Prior antibiotics**[e] | | | | | | |
| No | 41 | Ref | | 38 | Ref | |
| Yes | 182 | 0.83 (0.41-1.72) | 0.600 | 158 | 0.74 (0.32-1.72) | 0.500 |
| **Exclusive breastfeeding** | | | | | | |
| No | 334 | Ref | | 112 | Ref | |
| Yes | 94 | 0.90 (0.54-1.48) | 0.700 | 84 | 1.0 (0.48-2.11) | >0.900 |
| **Central line** | | | | | | |
| No | 405 | Ref | | 175 | Ref | |
| Yes | 23 | 1.51 (0.61-3.53) | 0.400 | 21 | 2.3 (0.69-8.01) | 0.200 |
| **Umbilical line** | | | | | | |
| No | 388 | Ref | | 165 | Ref | |

| | | | | | | |
|---|---|---|---|---|---|---|
| Yes | 40 | 2.02 (1.03-3.90) | 0.037 | 31 | 1.23 (0.47-3.22) | 0.700 |
| **Peripheral line** | | | | | | |
| No | 248 | Ref | | 47 | Ref | |
| Yes | 180 | 0.97 (0.64-1.47) | 0.900 | 149 | 1.40 (0.58-3.54) | 0.500 |
| **Intubation** | | | | | | |
| No | 370 | Ref | | 155 | Ref | |
| Yes | 58 | 1.76 (0.99-3.09) | 0.052 | 41 | 1.81 (0.78-4.27) | 0.200 |

a: OR = odds ratio; b: CI = confidence interval; c: aOR = adjusted odds ration; d: Weight measured on specimen collection; e: Antibiotics in the last two weeks

**Hospital A**

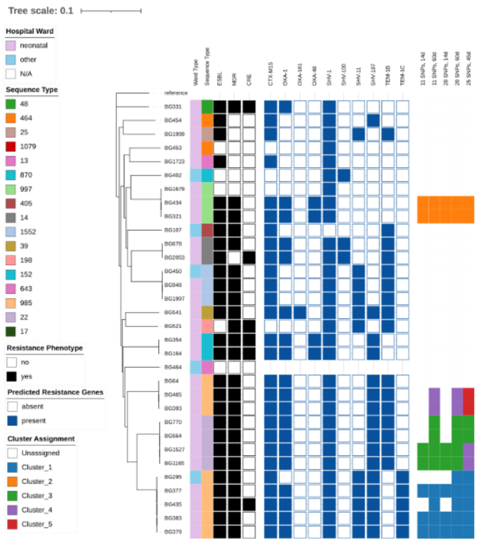


**Hospital B**

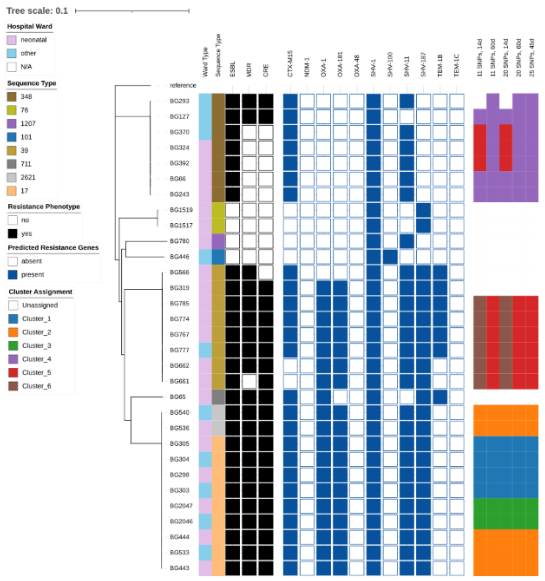


**Hospital D**

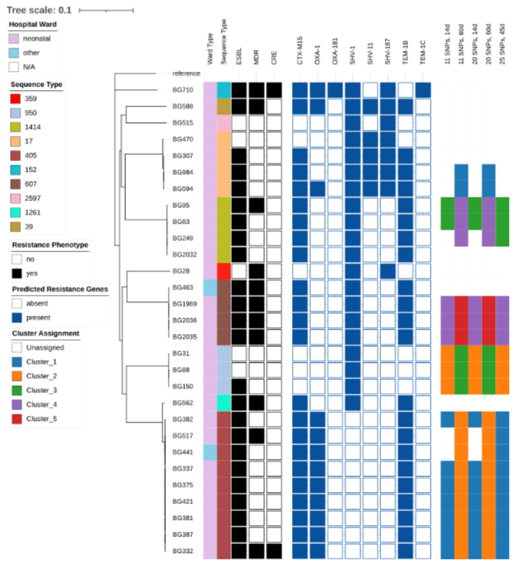


**Hospital C**

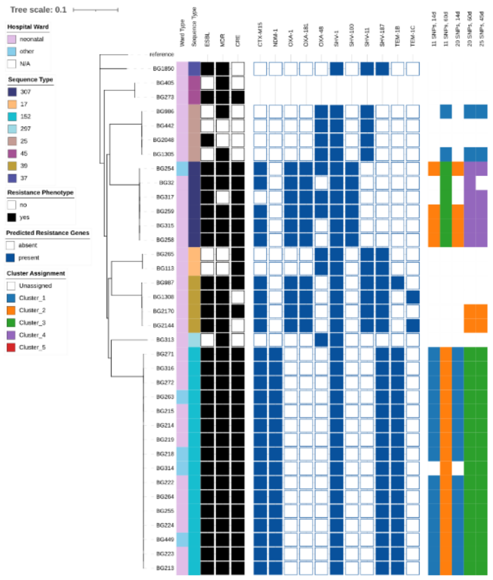


**Hospital E**

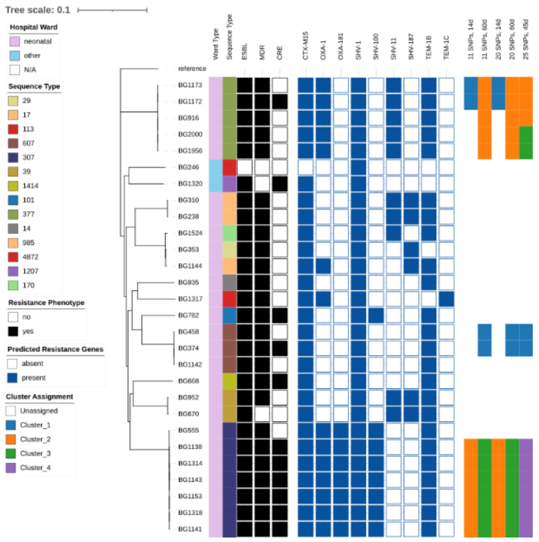


**Hospital F**

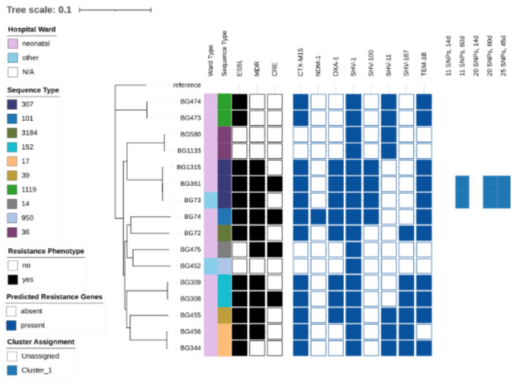

**Fig S2:** Phylogenetic trees showing selected phenotypic resistance and predicted genotypic resistance in *Klebsiella pneumoniae* isolates from case-patients at Hospital A-F.

Key: ESBL = extended-spectrum beta-lactamase producing (defined as resistance to one or more 3rd generation cephalosporin antibiotics); MDR = multi-drug resistant (defined as resistance to three or more antibiotic classes); CRE = carbapenem-resistant Enterobacterales (defined as resistance to one or more carbapenem antibiotics).

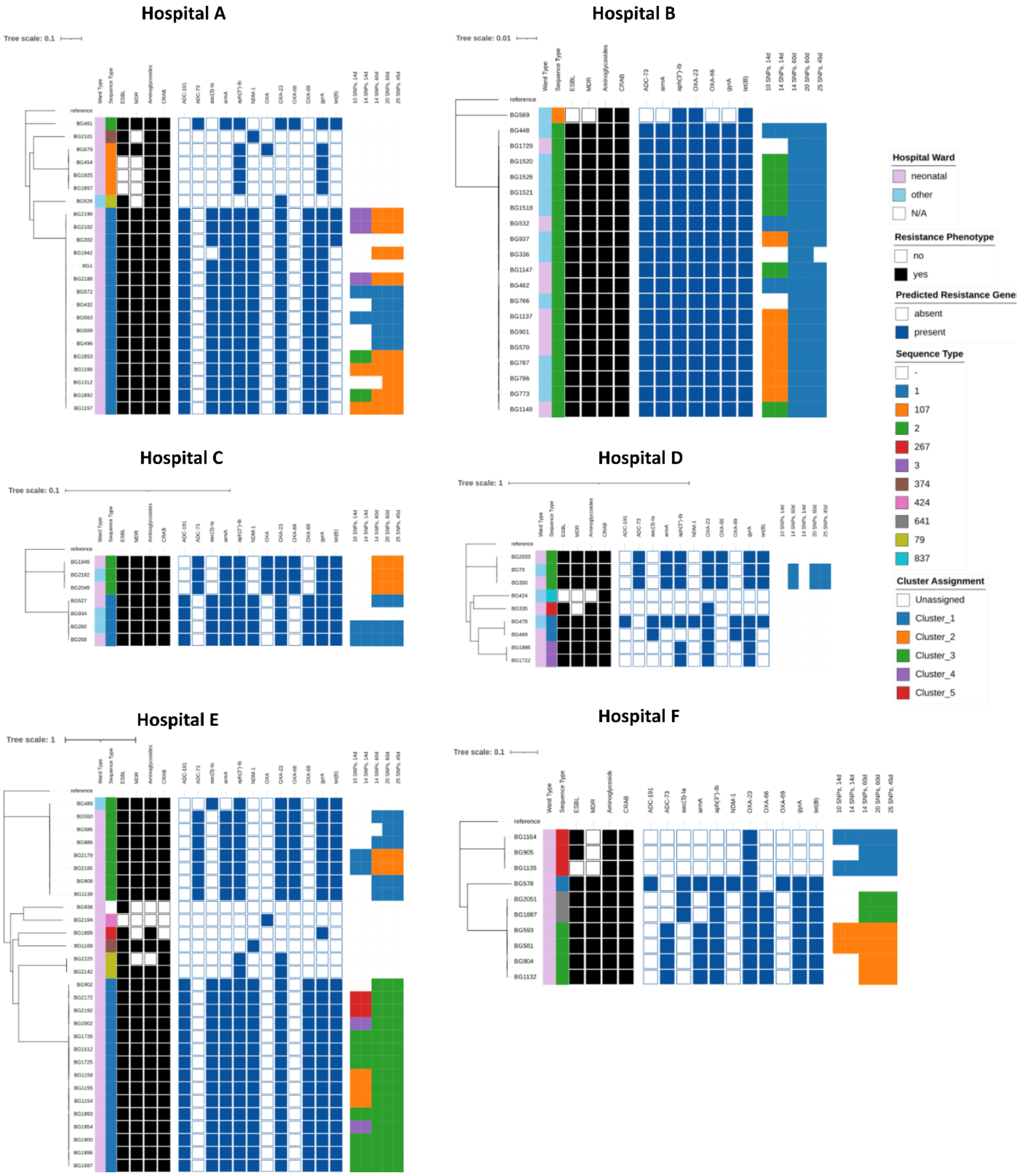


**Fig S3:** Phylogenetic trees showing selected phenotypic resistance and predicted genotypic resistance in *Acinetobacter baumannii* isolates from case-patients at Hospital A-F.

Key: ESBL = extended-spectrum beta-lactamase producing (defined as resistance to one or more 3rd generation cephalosporin antibiotics); MDR = multi-drug resistant (defined as resistance to three or more antibiotic classes); CRAB = carbapenem-resistant *Acinetobacter baumannii* (defined as resistance to one or more carbapenem antibiotics).

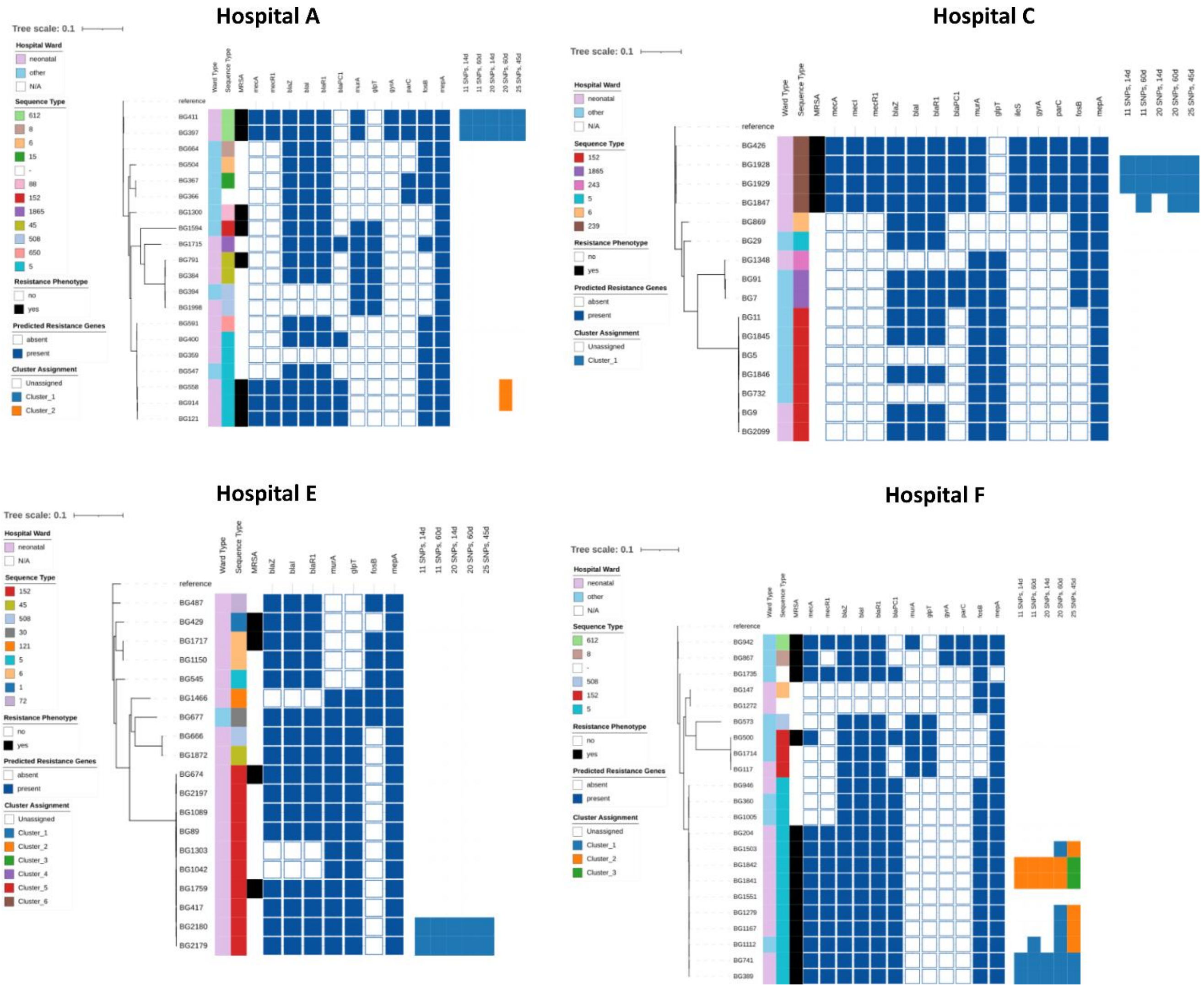


**Fig S4:** Phylogenetic trees showing selected phenotypic resistance and predicted genotypic resistance in *Staphylococcus aureus* isolates from case-patients at Hospital A, C, E and F. Key: MRSA = methicillin-resistant *Staphylococcus aureus* (defined as resistance to oxacillin or cefoxitin).

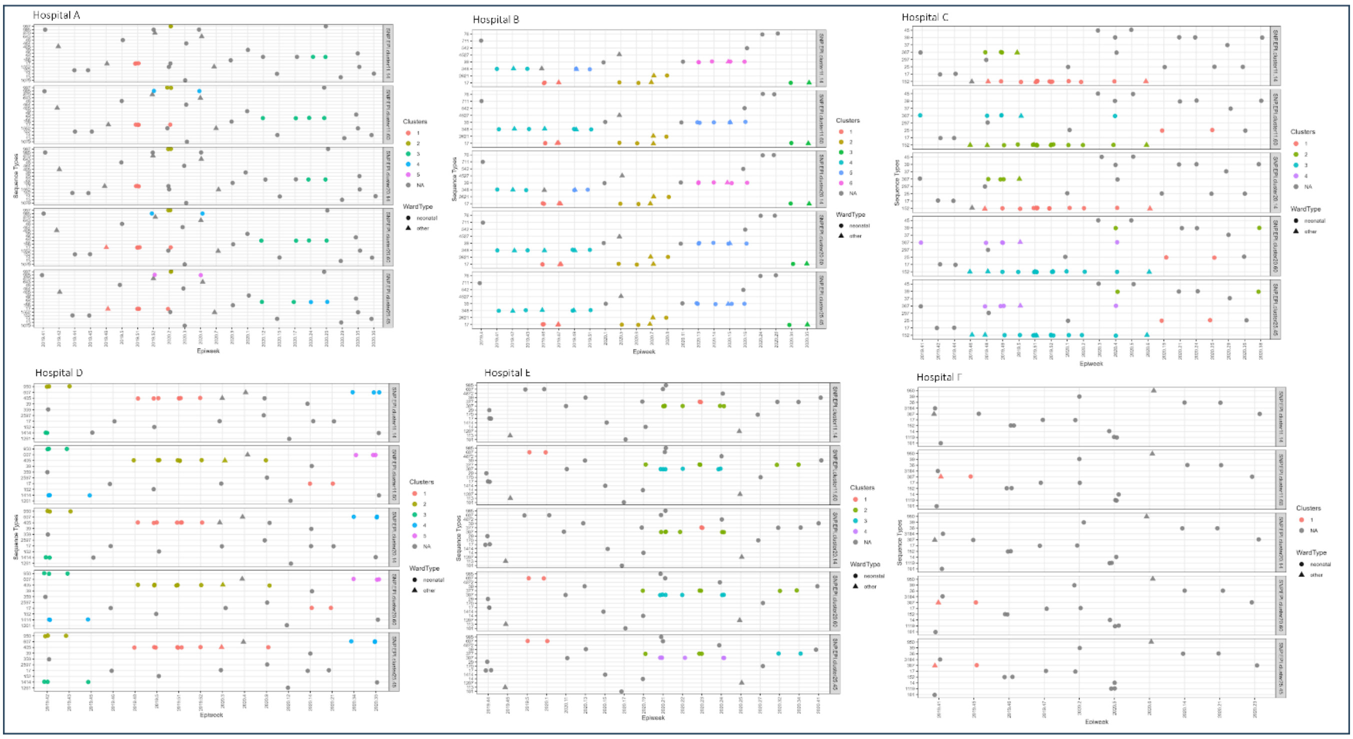


**Fig S5:** *K. pneumoniae* infection clusters detected in Hospital A to Hospital F by varying SNP-EPI criteria

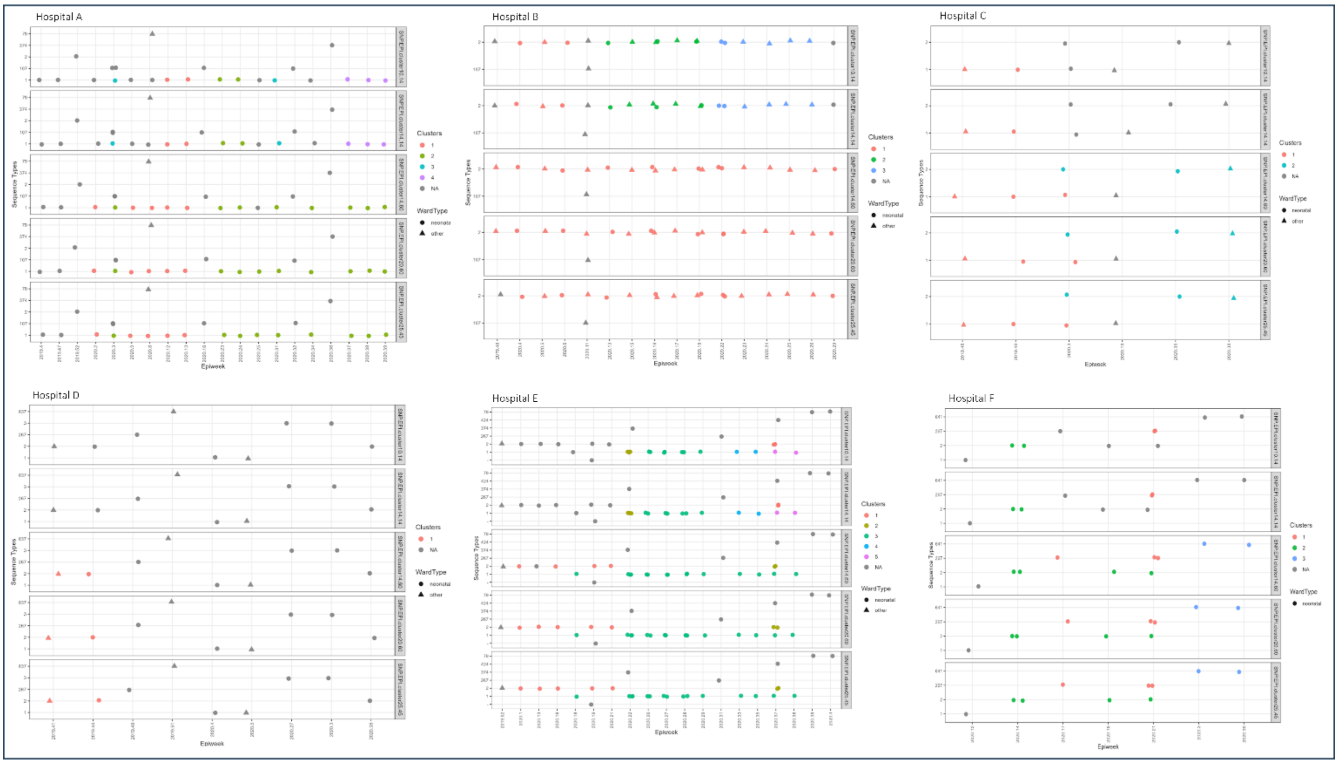


**Fig S6** *A. baumannii* infection clusters detected in Hospital A to Hospital F by varying SNP-EPI criteria

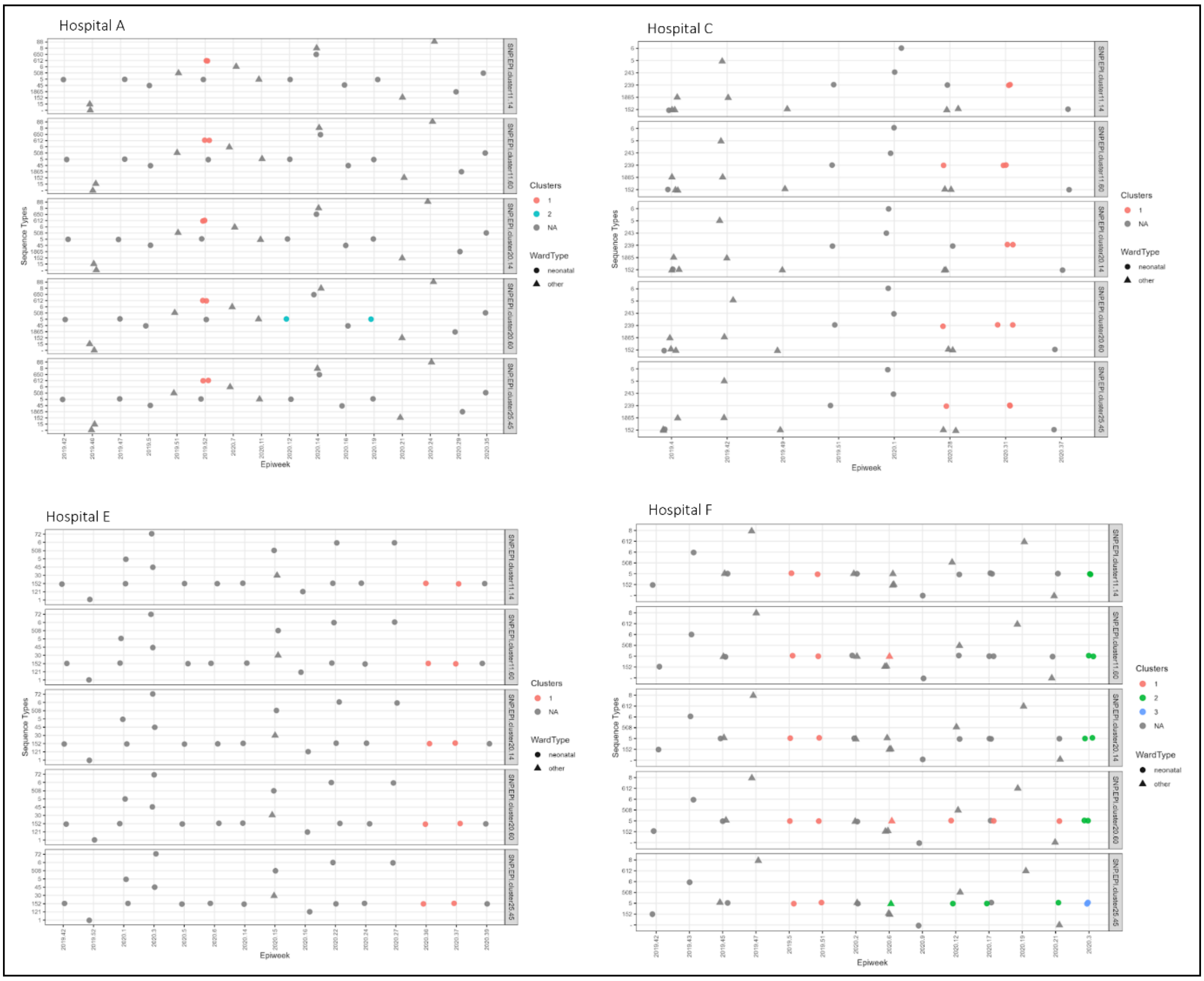


**Fig S7:** *S. aureus* infection clusters detected in Hospital A to Hospital F by varying SNP-EPI criteria
